# Integrated biological and chemical wastewater surveillance reveals local changes in dynamics of human-associated virus, bacteria and chemicals

**DOI:** 10.1101/2025.10.08.25337484

**Authors:** Anna J. Székely, Dhananjay Mukhedkar, Joakim Dillner, Ville N. Pimenoff

**Author notes:** Corresponding author: Ville Pimenoff PhD, University of Oulu.

## Abstract

Systematic analysis of environmental exposures (“exposomics”) has become a major research focus in recent years, as it will enable comprehensive studies of the effect of changing environmental exposures on human health over time and between communities. Wastewater-based surveillance (WBS) is a well-established tool for community surveillance of pathogens and chemicals. During and after COVID-19 pandemic between July 2021 and May 2024 we systematically collected wastewater samples at 18 wastewater treatment plants covering 15 cities in Sweden and encompassing 41% of the national population. The sample set consisted of different seasons, different geographical areas with different climates ranging from sub-artic to temperate oceanic, and different classes of cities ranging from metropolitan to commuting cities. Wastewater samples were concentrated and biotic profiling analyzed using shotgun metagenomics and abiotic profiling using LC-MS/MS.

From wastewater samples, we identified both known and unknown microbes, including human related bacteria and viruses, as well as sequences of unknown viral origin and a range of known and unknown environmental chemicals. We report a systematic detection of viral human pathogens and phages infecting bacteria typically found in human gut microbiota. Regional differences for both microbial and chemical exposome were identified. For two cities (Kalmar and Uppsala), we performed longitudinal sampling between 2022 and 2024 and found a high correlation of the bacterial, viral and chemical exposome in serial samples from the same city. Notably, there was consistently a 1.5-fold higher abundance of viruses in the wastewater of the city of Kalmar compared to Uppsala (P_FDR_=0.000007). This 1.5-fold higher abundance sustained across multiple human health-related pathogen species such as *JC polyomavirus*, gut bacteria infecting *Carrus communis* phages and *Enterococcus faecalis* bacteria. Seasonal changes in viral abundance in wastewater were also found (P_FDR_=0.001). Interestingly, we could also identify regional outbreaks of human infecting bacteria and virus such as Adenovirus F41 in Kalmar or changes in chemical abundance related to human activities and associated with the time of social relaxation of COVID-19 pandemic restrictions. In summary, systematic and integrated viral, bacterial, and chemical wastewater exposomics is a powerful tool for mapping of cumulative changes in human environmental exposures including unknown biological and chemical components and local outbreak features.

## Introduction

Wastewater-based surveillance (WBS) has a long research tradition. One of its first implementations was in Finland in the early 1960s, tracking poliovirus circulation to assess the efficiency of the national poliovirus vaccination program (Lapinleimu 1984; Harvala et al. 2021). While such early applications already demonstrated their value and significance, interest in wastewater monitoring has resurged globally in recent years due to the COVID-19 pandemic, which led to the rapid establishment of wastewater surveillance programs for SARS-CoV-2 and other emerging pathogens. This renewed attention has highlighted the broader potential of WBS as a powerful tool for mapping of the human exposome.

Most WBS programs currently focus on specific chemicals for monitoring environmental toxins or illicit drug use or on pathogen surveillance using qPCR, as demonstrated by the correlation between SARS-CoV-2 RNA levels in wastewater and COVID-19-related hospitalizations in the corresponding communities (Ahmed et al. 2020; Wade et al. 2022). The recent technological improvements in methodology, throughput, and price have enabled untargeted chemical profiling of thousands of chemicals using mass spectrometry and shotgun metagenome sequencing of more than tens of millions of reads from low biomass wastewater samples, enabling broader human biological exposome detection. These approaches allow the exploration of urban microbial ecosystems along with the chemical environment and simultaneously, enables the identification of both human-associated pathogenic microorganisms and chemicals related to human activities. These advancements in precision for monitoring biotic and abiotic exposures within WBS ultimately enables the estimation of total cumulative long-term exposures that likely affect our health.

Integrated Human exposomics aims to define the totality of environmental exposures that humans may encounter during lifetime, and includes measuring external factors such as pollutants, diet, lifestyle, microbes, occupational exposures, and social influences. This concept extends beyond traditional epidemiology to consider how these diverse exposures impact human health and disease risk. By integrating data from various sources, such as biomonitoring, environmental monitoring, and personal assessments, researchers aim to better understand the complex interactions between environmental factors and biological responses. Ultimately, the exposomics approach facilitates the identification of associations between exposure patterns and health outcomes, which can guide strategies for disease prevention and public health. Longitudinal sampling in WBS could be an important tool to assist in mapping the total environmental exposures at the community level.

In this study, we systematically collected wastewater samples from 15 cities across Sweden from 2021 to 2024. The sampling used here covers four full years and fifteen cities, ranging from the southernmost parts of Sweden to the far north. We employed shotgun sequencing on the wastewater samples and estimated the DNA exposome profiles in time and place in order to evaluate different sampling strategies on wastewater-based exposome profiling utilizing a comprehensive metagenomics algorithm.

## Materials and Methods

### Wastewater sampling

Between week 30, 2021 and week 21, 2024, an extensive collection of sewage samples (> 1000) was collected from Swedish municipalities to assess the SARS-CoV-2 pandemic. In this study, 91 sewage samples were selected from these samples representing various urban and rural locations in Sweden. The dataset encompasses both single-time and pooled water samples, with contributions from 15 municipalities managed by eighteen different water treatment companies (Table 1). The sampling timeline spanned across different weeks and years, offering a temporal view of sewage water characteristics. Notably, the largest longitudinal follow-up series were from Uppsala (N=23), Malmö (N=11) and Kalmar (N=15). After total nucleic acid extraction, the samples were aliquoted and stored at −80°C freezer.

**Table 1.**
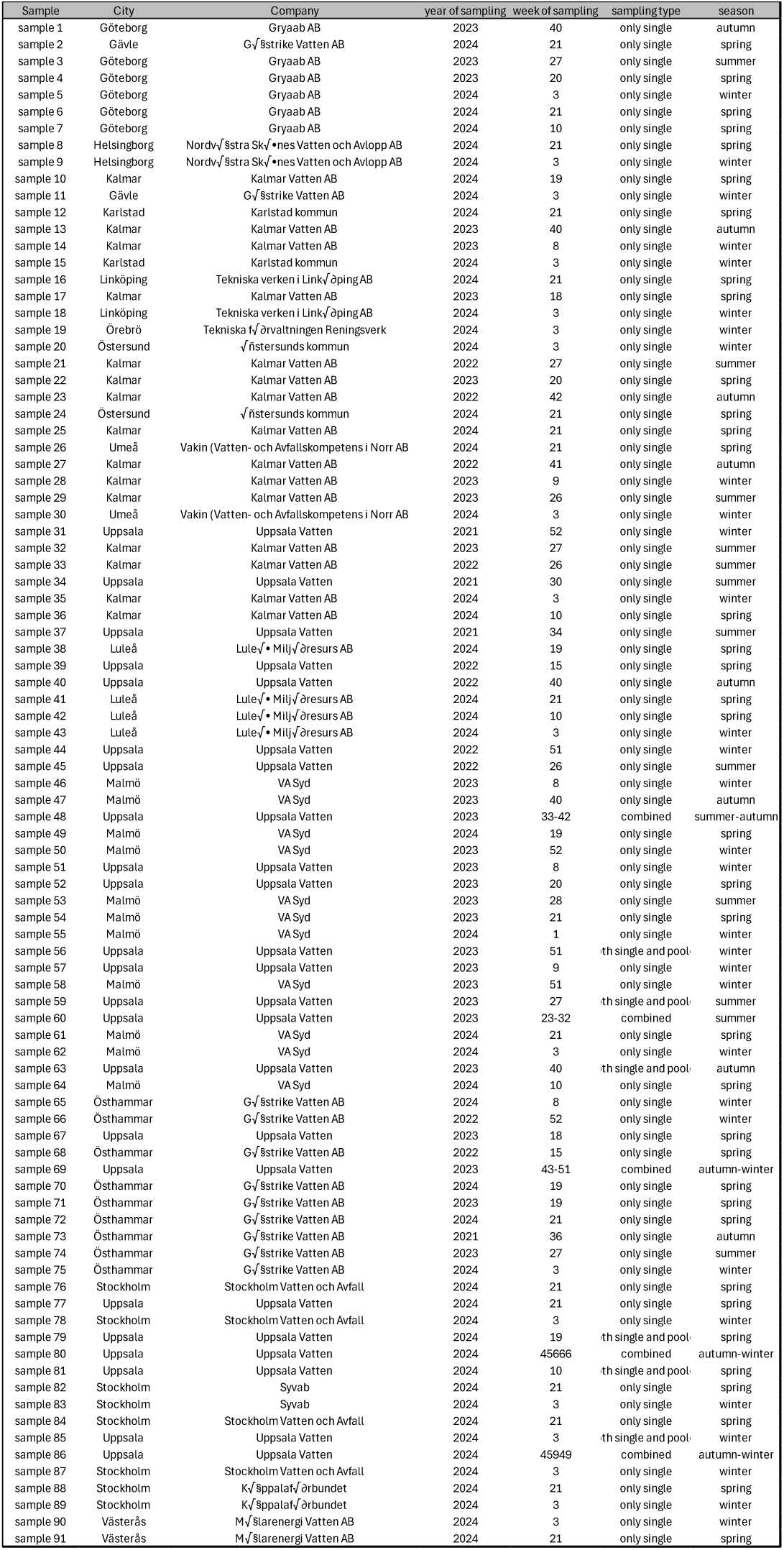
Wastewater sample characteristics.

### Wastewater biomass extraction and sequencing

Total genomic DNA was purified from 40 mL of sewage water samples. DNA extractions were performed using a Maxwell® RSC Enviro TNA Kit (Promega) for sewage samples, following the manufacturer’s instructions and eluted in 100 µL of nucleic acid-free water. Total genomic DNA was quantified using Qubit (ThermoFisher Scientific) measurements. Shotgun metagenomic sequencing was performed for all samples at the Finnish Functional Genomics Centre (FFGC, Finland). The quality of sample DNA was ensured using Agilent Bioanalyzer 2100 or Advanced Analytical Fragment Analyzer. Sample concentration was re-measured to ensure DNA quantity. Sixty nanograms of DNA were used for the library preparation using the *Illumina DNA Prep Library Preparation, Tagmentation kit (Illumina)* according to the library preparation protocol. The libraries were sequenced by the paired-end method (2 x 150 bp) on the Novaseq 6000 S4 platform.

### Taxonomic classification

The original paired-end metagenome sequences, obtained as fastq files, were quality-filtered using Trimmomatic and BBmap, including the removal of duplicate reads. Only high-quality reads (Q20) were retained for the microbial analysis. The sewage sample nucleic acid taxonomic classification was performed using a Kmer matching algorithm and a K-mer matrix from the fully curated archaea, bacteria, virus and eukaryotic pathogen genomes reference genome database (v.2025), including the NCBI Viral Genome resource (https://ncbiinsights.ncbi.nlm.nih.gov/2023/10/19/changes-virus-data-resources-ncbi/, accessed on 1 January 2025) and the Eukaryotic Pathogen Genome Database (http://www.eupathdb.org/, accessed on 1 January 2025). The classified species taxa were evaluated based on their at least 2% mapping coverage across the reference genome using our in-house pipeline and threshold to exclude potential false-positive taxa (available in GitHub: PimenoffV). In addition, we performed validation of the classification results against kraken2 classification (plusPFP reference genomes July, 2025). Taxonomic distribution and compositional analyses were conducted based on the observed read counts for each taxon, with various normalizing data transformations applied, as described in each analysis section.

### Extraction of wastewater chemicals

Frozen wastewater sample chemical extractions were performed in 20 sample batches. Samples were thawed overnight at 4°C and further to room temperature. Aliquots of 2 mL of wastewater were transferred to a furnaced glass tube, diluted (1:10) with 18mL LC-MS grade water (Optima, Fisher Chemical) and spiked with a mixture of isotope labelled internal standards (IS) to reach final concentration of 200 ng/mL. First, samples were vortexed and filtered using a 0.2 μm syringe filter of regenerated cellulose (Sartorius Minisart RC 15 mm). Subsequently, 10 mL of the sample were spiked with a volumetric IS (diuron-d6) and transferred to an injection vial for LC-HRMS analysis. For each batch three method blanks (LC-MS water Optima grade) were prepared in the same way in parallel with the samples. During sample preparation and extraction, samples and method blanks were prepared in a randomized order. Per each batch, a quality control (QC) sample was also prepared by pooling equal volumes (7 mL) of all prepared samples (excluding blanks).

### LC-HRMS analyses

In this study we used the non-targeted LC-HRMS chemical analysis method (SciLife, Sweden). Instrumental analysis was conducted by online SPE (Hypersil Gold ^TM^ PFP, 175 Å, 12 μm, 2.1 × 20 mm, and Hypersil Gold ^TM^ aQ, 175 Å, 12 μm, 2.1 × 20 mm; Thermo Scientific ^TM^) followed by analytical separation on a reversed phase column (Acquity UPLC BEH C18, 1.7 μm, 2.1 × 100 mm, 130 Å; with an Acquity UPLC BEH C18 VanGuard Precolumn 1.7 μm, 2.1 × 5 mm, 130 Å; Waters) at 40°C. Sample injections (1 mL, full loop, 130 s loading time at 0.5 mL/min) were performed in randomized order. Injections of QC samples were made at the beginning of each sequence, and after every 5-7 sample injections.

Data acquisition was performed in two ionization modes (ESI+, ESI-) on a QExactive Orbitrap HF-X (Thermo Fisher Scientific) operated with H-ESI spray capillary voltages were +3 kV (ESI+) and −2.5 kV (ESI-), with capillary (ion transfer tube) temperature of 300°C, sheath gas flow rate set to 50, auxiliary gas to 16, sweep gas to 2, and an auxiliary gas temperature of 400°C. The AGC target in both full scan and DIA method was set to 1e6. Spectral acquisition alternated between full scan MS1 (*m/z* 80-1000, nominal resolution 120,000 at *m/z* 200) and nontarget data-independent MS2 acquisition (DIA) using five sequential precursor isolation windows (i.e. *m/z* 70-140, 135-205, 200-300, 190-505, 500-1000) at nominal resolution of 30,000, RF lens set at 50%, and normalized HCD collision energy (20, 70 V), with fragments formed at different collision energies analysed simultaneously and averaged in the Orbitrap mass analyser.

### Data processing and spectral library annotations

Raw mass spectrometry data was pre-processed using the open source software MS-DIAL^3^ (v. 4.9.221218; https://systemsomicslab.github.io/compms/msdial/main.html) for chromatographic alignment across samples, peak area integration, DIA spectral deconvolution, and extraction of feature lists with accurate masses. Features lists were exported from MS-DIAL by filtering with criteria that at least one sample had a peak area 10 times higher than the average of all method blanks (n=15).

Data were normalized as previously described^4^ using internal standards (IS) and applying a multivariate PCA (UVN scaling, PC1) to model IS response variation across samples, whereby PC1 scores were used to normalize all features in each sample in the dataset. Feature areas were subsequently blank-subtracted by removing the mean area of method blanks. Additional cleanup included removal of redundant adducts and in-source fragments. Annotations of LC-HRMS features were performed by spectral matching to open-access libraries MassBank Europe (https://massbank.eu/), MassBank North America (https://mona.fiehnlab.ucdavis.edu/), and GNPS (https://gnps.ucsd.edu/). The annotations were reported with confidence Level 2a (Schymanski *et al*.,^5^) and considering an accurate MS mass deviation of maximum 0.002 Da between the detected precursor ion and the library record, and a MS spectral matching with a total identification score assigned by MS-DIAL of > 700 (“total score”).

## Results

A total of 91 wastewater samples collected between July 26^th^ 2021 and May 26^th^ 2024, were selected and retrospectively analyzed using shotgun metagenomics for known domains of life (archaea, bacteria, and eukarya) as well as the presence of known and currently unknown viral species taxa. Pre-processed and quality-curated metagenomes from the 91 wastewater samples comprised between 20.6 million and 193.7 million high-quality 150 bp PE sequences per sample. The wastewater samples originated from 15 Swedish cities, namely Göteborg, Helsingborg, Kalmar, Luleå, Malmö, Östhammar, Stockholm, Gävle, Karlstad, Linköping, Örebro, Östersund, Umeå, Uppsala and Västerås (Fig. 1). Sample set included both single time (N=86) and cumulative pooled water samples (N=5) to test if cumulative exposure estimation is possible by sample pooling (Table 1). A subset of the samples represents longitudinal wastewater sampling serieses from the municipalities of Uppsala (N=23), Kalmar (N=15), and Malmö (N=11) (Table 1).

**Figure 1.**
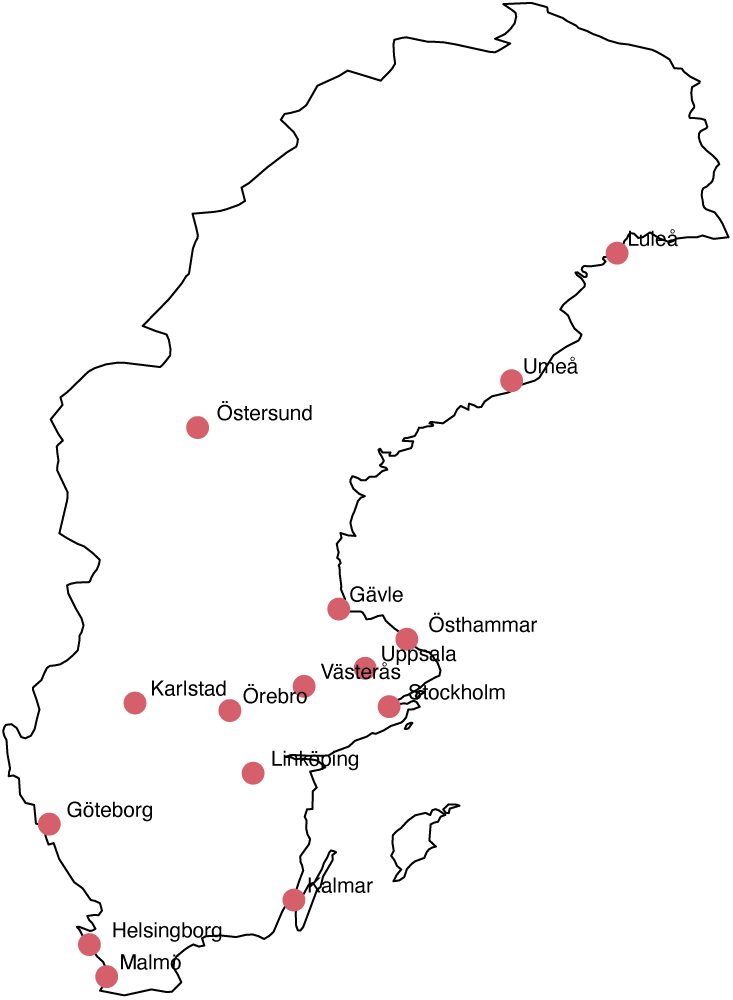
Sampling. All 15 wastewater sites sampled in this study.

### Biotic compounds in wastewater

Our metagenomic classification analysis here targeted archaea, bacteria, viruses and eukaryotic pathogen genomes and revealed a significant fraction of archaea, bacteria and viruses in the wastewater samples and human DNA as eukarya (Fig. 2A). Bacteria were the most dominant taxa classified across the wastewater samples representing more than 95% of the assigned taxa across the metagenomes. In comparison, viruses represented at least 2.5% and archaea at least 0.1% of the classified sequences in most wastewater metagenomes (Fig. 2A). Bacteria were detected at similar relative abundance across the studied municipalities. In contrast, specific anomalies of high archaea (i.e. Stockholm and Umeå) and viral (i.e. Gävle, Kalmar and Stockholm/Henriksdahl) abundances were observed for a few cities (Fig. 2B).

**Figure 2.**
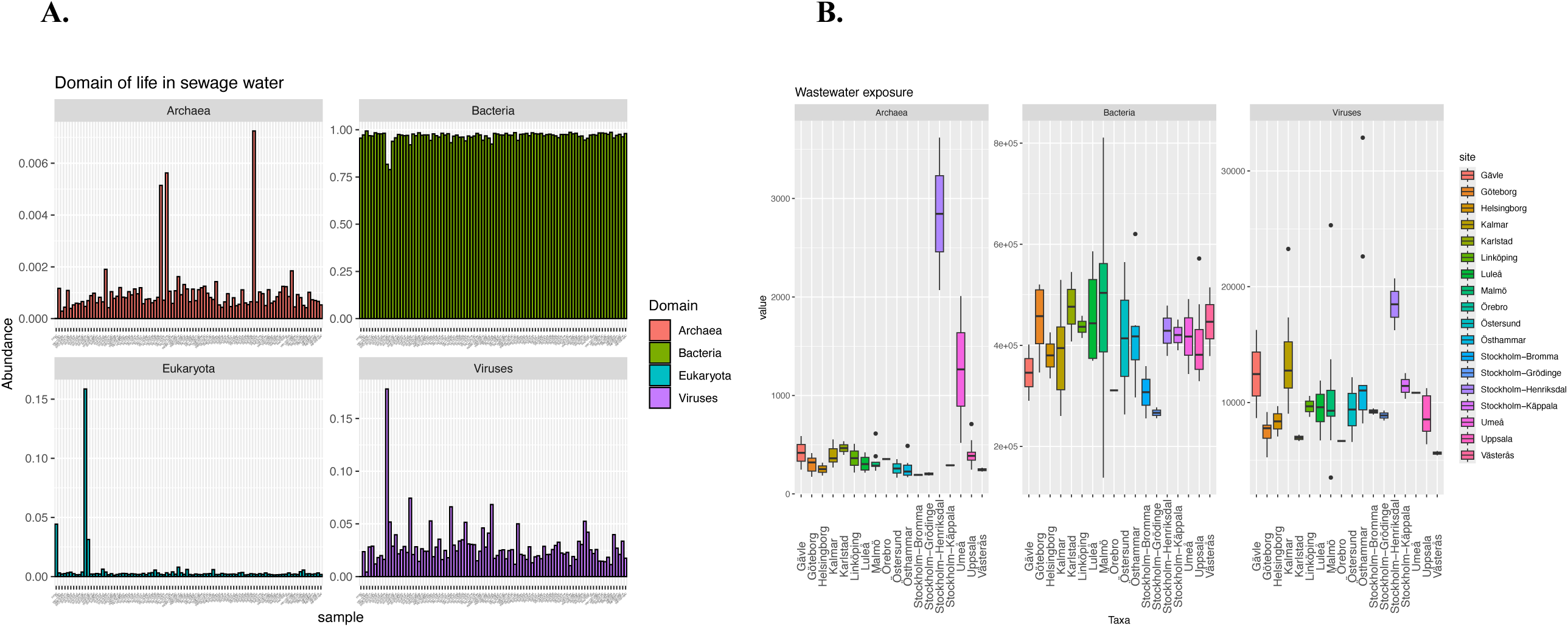
Domains of life observed in wastewater. **A.** Relative proportions of high-quality DNA reads classified into the four domains of life from wastewater samples. **B.** Domain-level relative abundances presented separately for each municipality.

To assess longitudinal trends in microbial community composition, we analyzed the relative abundances of archaea, bacteria, and viruses in wastewater using a four-week sliding window across the sampling period from 2021 to 2024 in Uppsala and Kalmar, the two municipalities with the longest follow-up (Fig. 3). The relative abundance of total viral content was significantly 1.5-fold higher in Kalmar compared to Uppsala during the period from autumn 2022 to spring 2024 (P_FDR_ = 0.000007), with a declining trend observed in both municipalities over time (Fig. 3A). In contrast, bacterial relative abundances did not differ significantly between Kalmar and Uppsala during the same period (P_FDR_ = 0.46), but some temporal fluctuations were apparent in the sliding window analysis (Fig. 3B), as well as when stratified by sampling year or by sampling time.

**Figure 3.**
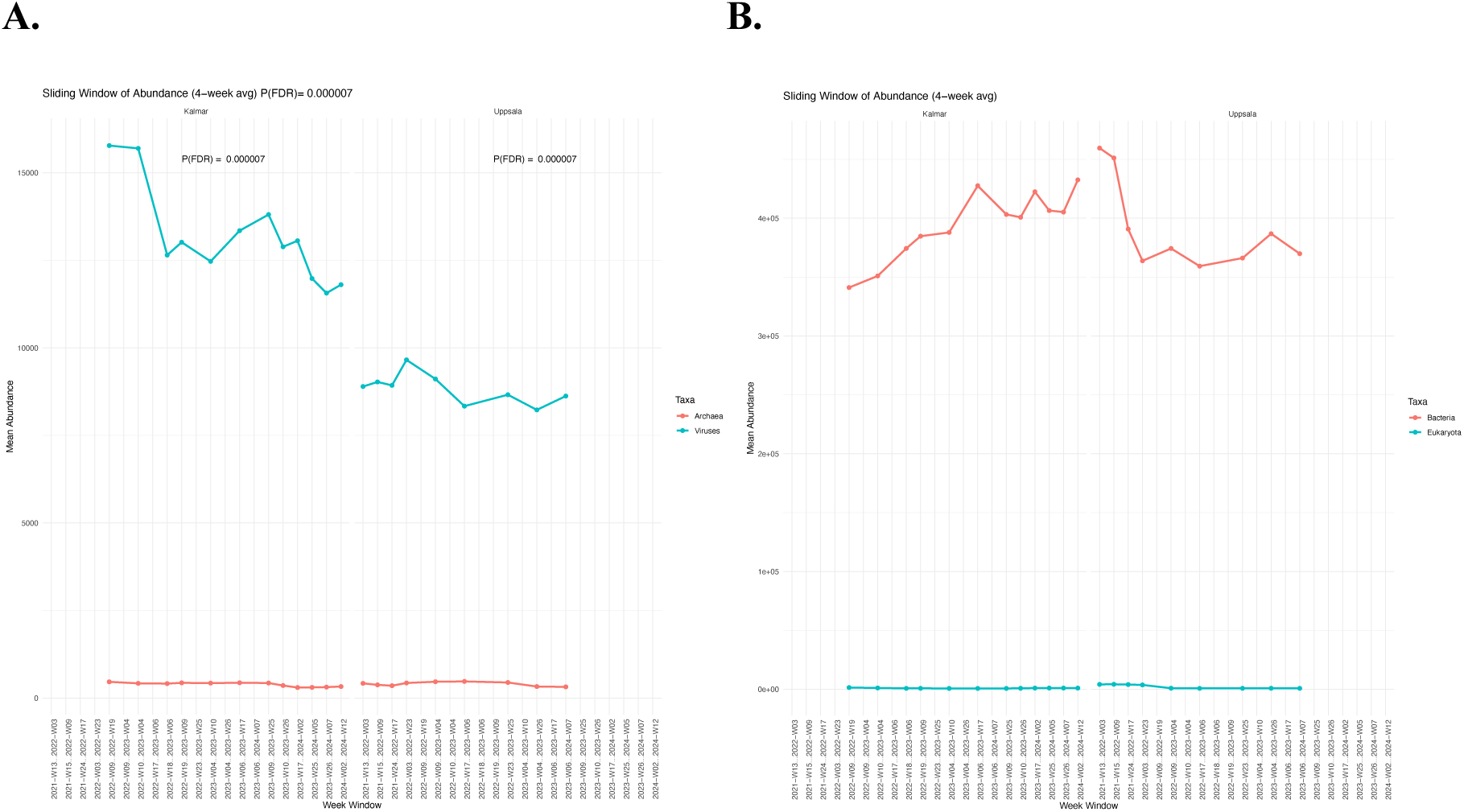
The relative abundance of A) all classified archaea and viruses and B) all bacteria and eukarya (B) in a sliding window of four weeks throughout the sampling from 2021 till 2024. Relative abundance log-tansformed and normalized with total number of high-quality reads.

### Human viral and bacterial pathogens in wastewater

The wastewater virome analysis revealed a total of 1,362 viral species with confidently detected genome coverage after correcting for false positives. Of these, 537 species accounted for more than 1% of the total classified viral taxa. Focusing on human viral pathogens within the wastewater virome, we identified six viral species or lineages known to infect humans and affect health (Table 2), which originate from nine eukaryotic human viral families: Adenoviridae, Hepatitisviridae, Herpesviridae, Papillomaviridae, Parvoviridae, Polyomaviridae, Poxviridae, and Reoviridae (Bai et al. 2022; Anon 2024). The six identified human infecting viral DNA lineages were *JC polyomavirus, Human Adenovirus F41, A61 and C12 serotypes*, *Molluscipoxvirus sp.* and *Human gammaherpesvirus 4* (Table 1).

**Table 2.** Prevalence of the most observed human and human gut bacteria infecting viruses across the wastewater samples.

| Viral lineage | Species | Host | Wastewater |  |
| --- | --- | --- | --- | --- |
|  |  |  | N | % |
| JC polyomavirus | Polyomavirus | Human | 91 | 100% |
| Human adenovirus 41 | Adenovirus F | Human | 85 | 93% |
| Human adenovirus 61 | Adenovirus A | Human | 33 | 36% |
| Molluscipoxvirus | Poxvirus | Human | 12 | 13% |
| Human adenovirus 12 | Adenovirus | Human | 3 | 3% |
| Human gammaherpesvirus 4 | Epstein-Barr virus | Human | 2 | 2% |
| Carjivirus communis | Phage | Gut bacteria | 91 | 100% |
| Carjivirus hominis | Phage | Gut bacteria | 91 | 100% |
| Endlipuvirus intestinihominis | Phage | Gut bacteria | 91 | 100% |
| Kingevirus communis | Phages | Gut bacteria | 91 | 100% |
| Septimatrevirus kakheti25 | Phage | Pseudomonas spp. | 91 | 100% |
| Tunggulvirus f2 | Phage | Pseudomonas spp. | 87 | 96% |
| Pseudomonas phage BUCT-F | Phage | Pseudomonas spp. | 38 | 42% |
| Pseudomonas phage Kaya | Phage | Pseudomonas spp. | 1 | 1% |
| Coopervirus nigel | Phage | Mycobacterium spp. | 6 | 7% |
| Coopervirus cooper | Phage | Mycobacterium spp. | 2 | 2% |
| Coopervirus vincenzo | Phage | Mycobacterium spp. | 2 | 2% |
| Coopervirus stinger | Phage | Mycobacterium spp. | 1 | 1% |
| Mushuvirus mushu | Phage | Faecalibacterium spp. | 91 | 100% |
| Toutatisvirus toutatis | Phage | Faecalibacterium spp. | 91 | 100% |
| Seoulvirus SPN3US | Phage | Salmonella phage | 91 | 100% |
| Popoffvirus pv56 | Phage | Fish infecting bacteria | 91 | 100% |
| Acinetobacter virus 133 | Phage | Acinetobacter spp. | 91 | 100% |
| Methanoculleus virus L4768 | Phage | Archaea | 39 | 43% |
| unclassified viruses | unknown | unknown | 91 | 100% |

Furthermore, an array of bacteriophages, viruses that infect bacteria, was detected in the wastewater samples. From these, we selected eighteen most abundant phages, representing nine distinct groups for further analysis: 1. human gut bacteria infecting prototypical *crAssphage (p-crAssphage) Carjivirus communis, Carjivirus hominis;* 2. human gut *Faecalibacteria* infecting crAss-like phages *Mushuvirus mushu* and *Toutatisvirus toutatis*; 3. Other human gut bacteria infecting phages *Endlipuvirus intestinihominis* and *Kingevirus communis*; 4. Pseudomonas bacteria infecting phages *Septimatrevirus kakheti25, Tunggulvirus f2, Pseudomonas phage BUCT-PX-5* and *Pseudomonas phage Kaya*; 5. Mycobacteria infecting phages *Coopervirus nigel, Coopervirus cooper, Coopervirus Vincenzo and Coopervirus stinger*; and *6. Salmonella enterica* bacteria infecting phage *Seoulvirus SPN3US;* 7. fish bacteria infecting *Popoffvirus pv56 phage*; 8. Environmental Acinetobacteria infecting *Acinetobacter virus 133 phage* and 9. Archaea infecting *Methanoculleus virus L4768 phage* (Table 2). Moreover, we detected significant amounts of viral DNA of unclassified viruses in each wastewater sample of this study (Table 2).

After correcting for false positives, the wastewater bacteriome analysis revealed almost 6000 bacterial species from 401 genera with confidently detected genome coverage. Of these, we selected sixteen bacterial species or lineages known to be pathogenic to humans (Table 3). Ten bacterial species commonly linked to gastrointestinal infections and related conditions such as gastroenteritis and diarrhea were identified. These included *Clostridioides difficile*, *Salmonella enterica*, *Vibrio cholerae*, *Campylobacter jejuni*, *Shigella flexneri*, *Escherichia coli* O157:H7, *Escherichia coli* ETEC H10407, *Helicobacter pylori*, *Shigella dysenteriae* and *Shigella boydii* (Table 3). Six opportunistic bacterial pathogens commonly associated with infections such as pneumonia were identified, including *Acinetobacter baumannii*, *Enterococcus faecalis*, *Klebsiella pneumoniae*, *Mycobacterium avium* complex (MAC), the *Pseudomonas aeruginosa* group and *Staphylococcus aureus* (Table 3).

**Table 3.**
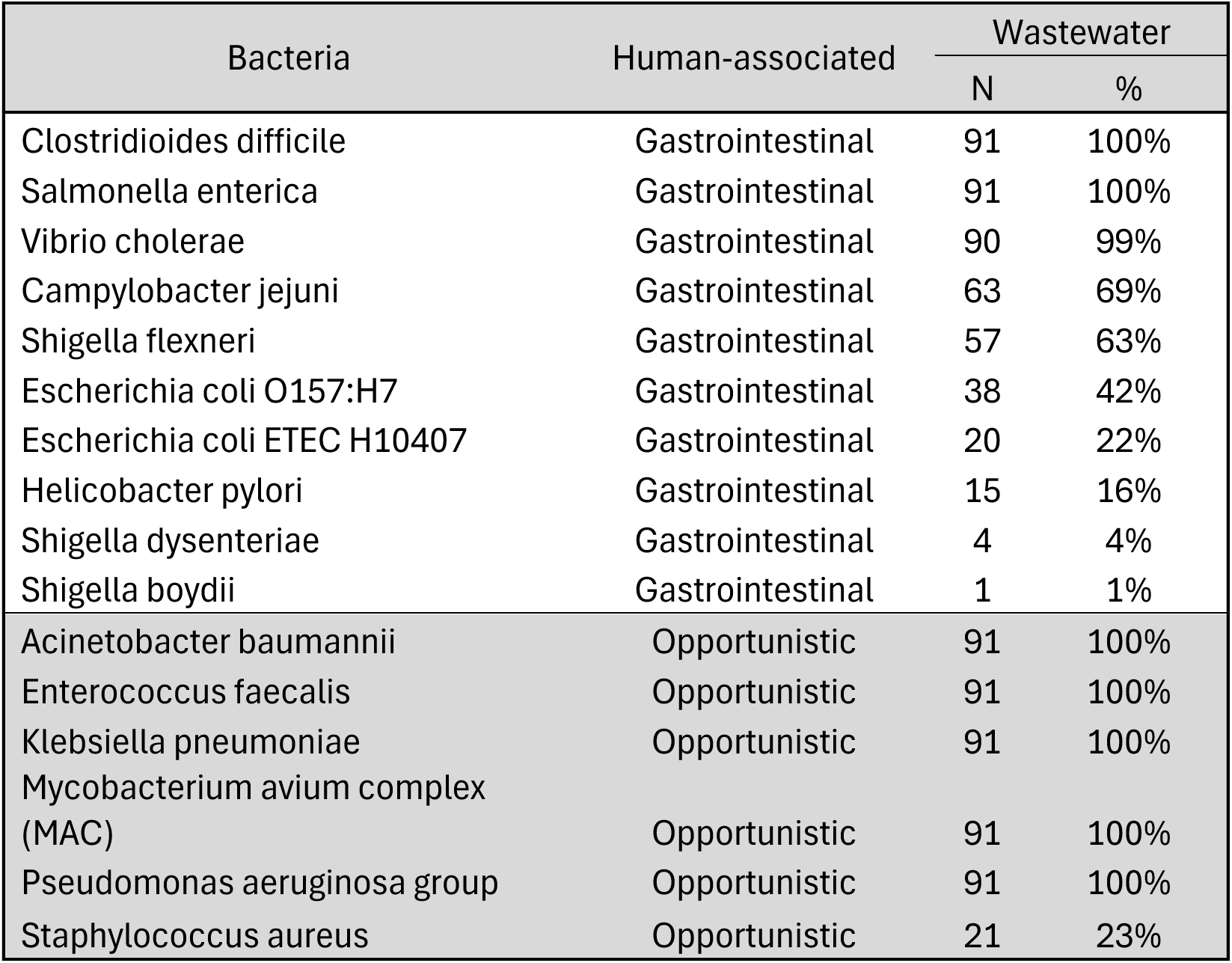
Prevalence of the most abundant human-associated bacterial pathogens across the wastewater samples.

### Regional and temporal exposure to pathogens

Ordination of the confidently detected 537 viral species and 401 bacterial genera both revealed significantly different (P = 0.001) community compositions between Kalmar, Malmö, and Uppsala wastewater systems (Fig. 4). A seasonal shift in wastewater microbes was also observed for all 537 classified viral species (P = 0.002) and the 401 bacterial genera (P=0.007). Still, this shift was only significant in Kalmar wastewater (Fig. 5). In agreement, the selected twenty-four viral (and the proportion of unclassified viral sequences) and selected sixteen bacterial human-related pathogen lineages detected in wastewater from Kalmar, Malmö, and Uppsala showed significantly distinct (P=0.001) compositional profiles (Fig. 6), but the seasonal shift was significant (P=0.001) only in Kalmar.

**Figure 4.**
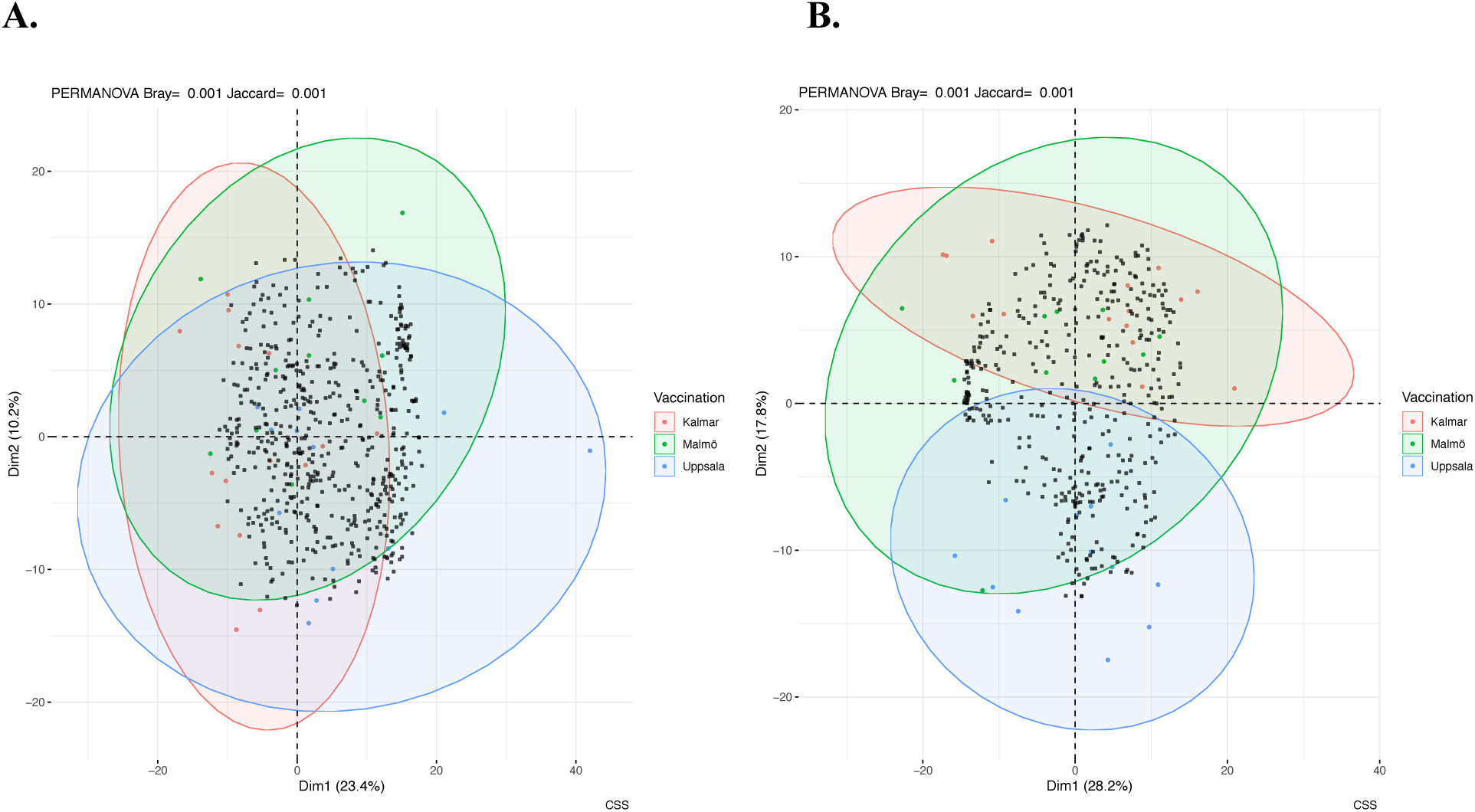
Ordination of all classified viral and bacterial species above 1% of the total abundance in wastewater. **A.** Viral community composition of 537 species taxa differed significantly (P = 0.001) between the cities of Kalmar, Malmö, and Uppsala. **B. Total of 398 bacterial genera revealed significantly different (P=0.001) community composition in wastewater** between the cities of Kalmar, Malmö, and Uppsala.

**Figure 5.**
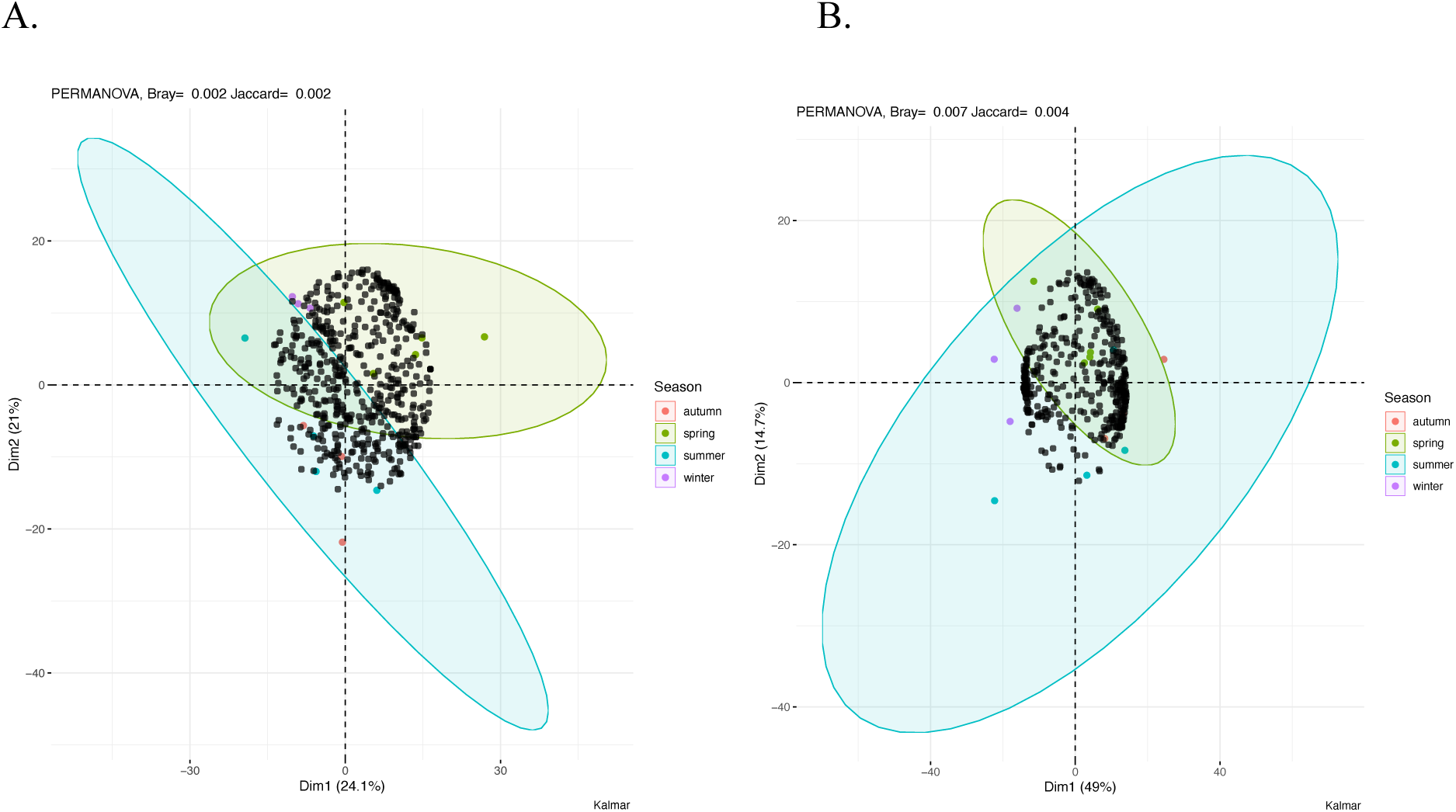
Significant seasonal variation in the composition of wastewater-associated virus and bacteria. **A.** Significant seasonal shift in viral community structure based on 537 viral taxa was observed in Kalmar wastewater (P = 0.002). **B.** Bacterial community composition of 401 genera showed a significant seasonal variation in Kalmar wastewater (P = 0.007).

**Figure 6.**
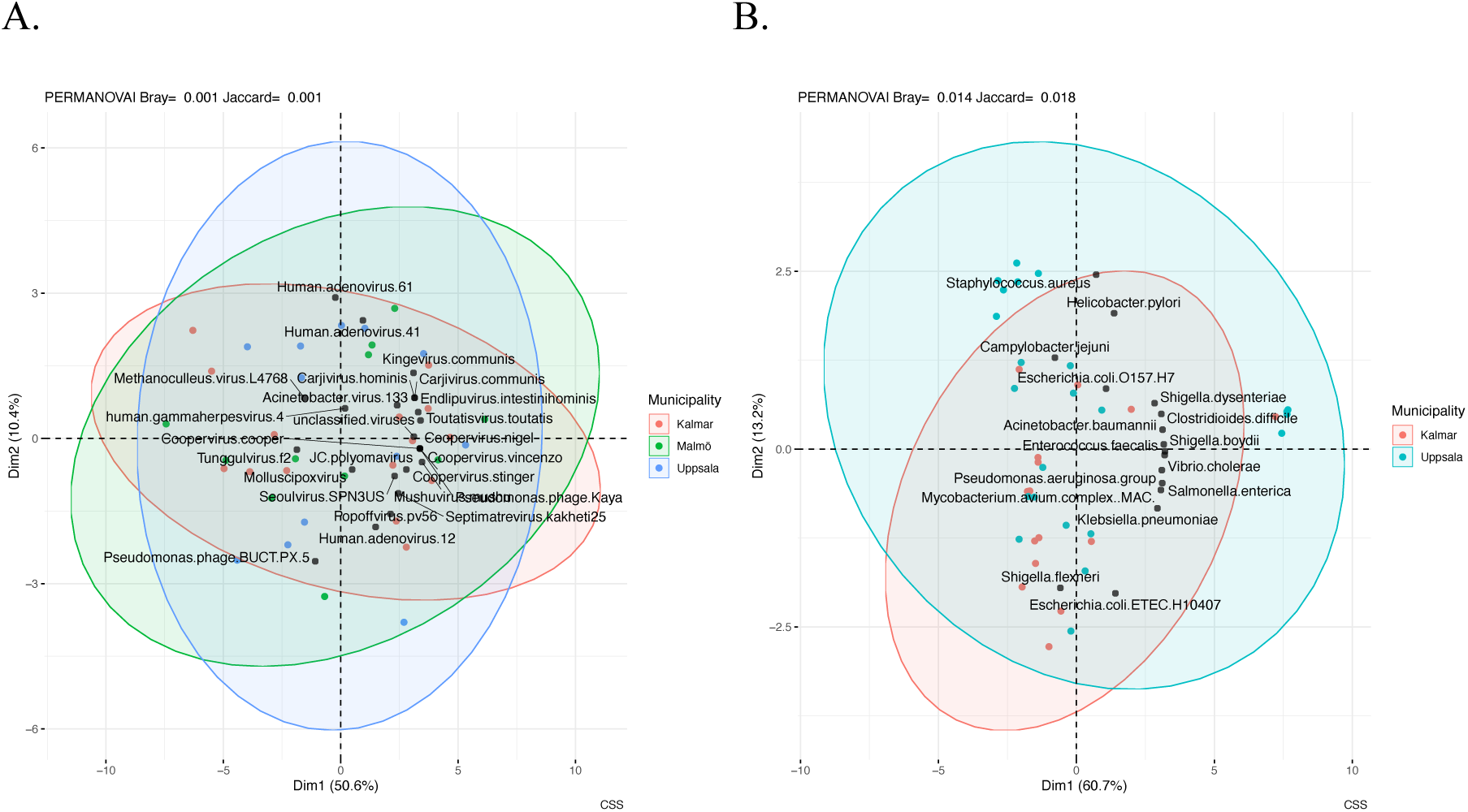
Ordination of selected human-related viral and bacterial taxa in wastewater between municipalities. **A.** The composition of selected 24 human-related viral taxa and the taxa of unclassified viruses showed significantly different compositional profiles between Kalmar, Malmö, and Uppsala (P = 0.001). **B.** The sixteen human-pathogenic bacterial species showed a significant compositional difference between Kalmar and Uppsala (P = 0.014).

From the most abundant human-related viruses and phages, we selected sixteen taxa observed in at least 36% of the wastewater samples for more in-depth regional and temporal analysis (Table 2). The human-infecting *JC polyomavirus* and *Human Adenovirus F41* lineages were observed in all and 93% of the wastewater samples, respectively, while *Human Adenovirus A61* was observed in 36% of the samples (Table 2). The regional prevalence estimate for each pathogen across the municipalities was mostly in the same range, with few cities showing significantly different relative abundances (Fig. 7A). Selecting the two municipalities with most of the follow-up samples a 1.5-fold significantly (P=0.0000068) higher accumulated *JC polyomavirus* relative abundances were observed in wastewater of the city of Kalmar compared to Uppsala (Fig. 6B) As a function of a four-week sliding window of sampling time, the relative abundance of *JC polyomavirus* is declining in Kalmar wastewater. In contrast, the relative abundance is lower and relatively stable in wastewater from Uppsala (Fig. 7B) In contrast, the four weeks sliding window revealed a rapid increase in relative abundance of *Adenovirus F41* from 2023 to 2024 in Kalmar wastewater, while a decrease in abundance was observed in Uppsala during the similar sampling frame (Fig. 7C)

**Figure 7.**
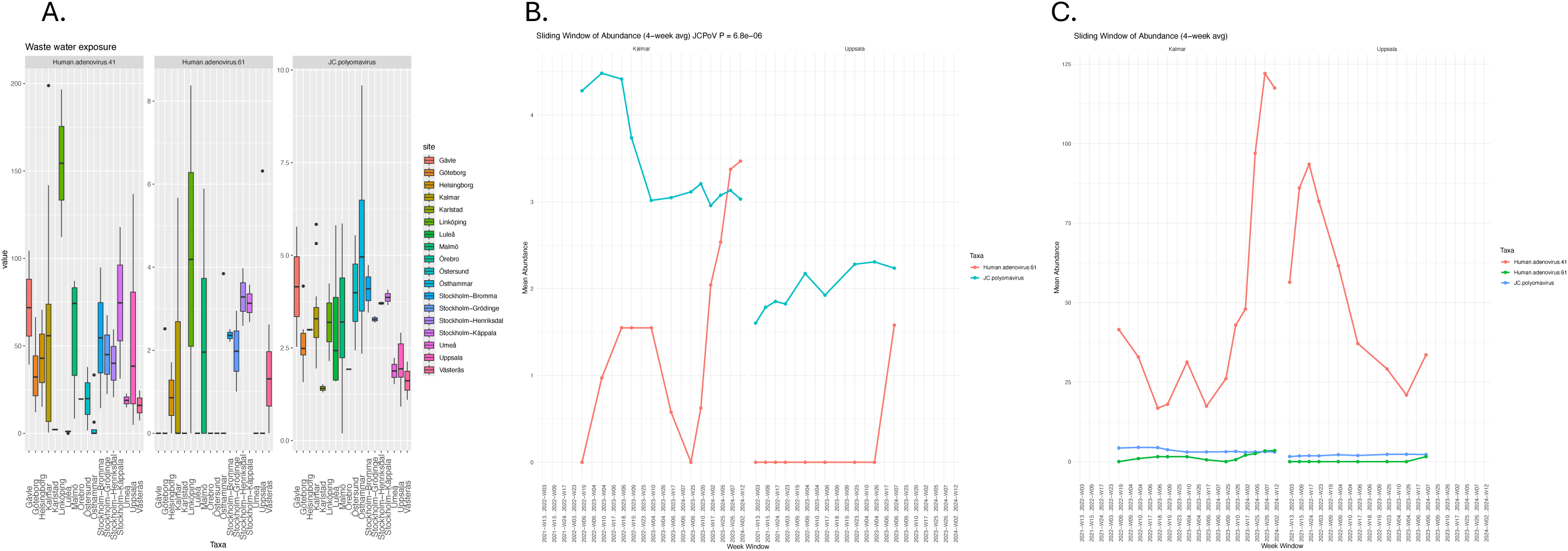
Most prevalent human-infecting viral pathogens detected in wastewater. **A.** JC polyomavirus and Adenovirus lineages F41 and A61 detected across 15 Swedish municipalities. **B.** Four-week sliding window analysis of JC polyomavirus and Adenovirus A61 relative abundance from 2021 to 2024. **C.** Four-week sliding window analysis of Adenovirus F41 and A61 relative abundance over the same period. All relative abundances were normalized with total number of high-quality reads and log-tansformed across the whole dataset.

The four human gut bacteria infecting phages *Carjivirus communis, Carjivirus hominis, Endlipuvirus intestinihominis* and *Kingevirus communis* were observed in all of the wastewater samples across the 15 Swedish municipalities (Table 2) with similar relative abundance (Fig 8A). Selecting the three municipalities with most of the follow-up samples, a significantly 1.5-fold higher relative abundance was systematically observed for all four phages in wastewater from Kalmar and Malmö compared to Uppsala (Fig. 8B) and a systematic trend of increased abundance for all phages in 2024 (Fig. 8C).

**Figure 8.**
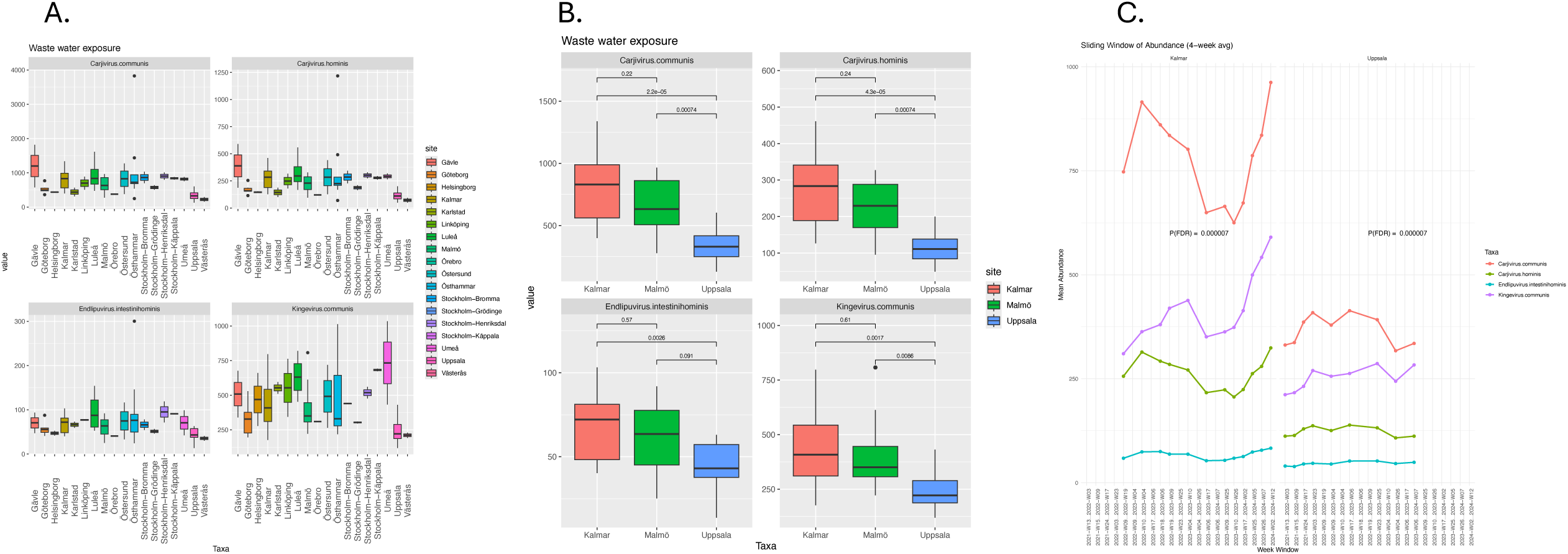
Four prevalent phages infecting human gut bacteria identified in wastewater. **A.** Presence of *Carjivirus communis*, *Carjivirus hominis*, *Endlipuvirus intestinihominis*, and *Kingevirus communis* across samples from 15 cities. **B.** Significantly higher relative abundances of these phages in wastewater from Kalmar and Malmö compared to Uppsala. **C.** Seasonal variation in relative phage abundance over a 4-week sliding window from summer 2021 to spring 2024 in Kalmar and Uppsala wastewater samples.

The three *Pseudomonas* spp. infecting phages (Table 2); *Septimatrevirus kakheti25* was observed in all samples, *Tunggulvirus f2* was observed in 96%, and *Pseudomonas phage BUCT-PX-*5 was observed in 42% of the wastewater samples with highly similar relative abundance across the 15 Swedish municipalities (Fig 9A). Selecting the municipalities with most of the follow-up samples, a significantly higher relative abundance was observed for *Tunggulvirus f2* and *Pseudomonas phage BUCT-PX-5* in wastewater from Uppsala compared to Kalmar and Malmö (Fig. 9B). No clear seasonal or regional pattern was observed for the three *Pseudomonas* spp. infecting phages, except that the *Tunggulvirus f2* phage was highly prevalent only in wastewater from Uppsala (Fig. 9C).

**Figure 9.**
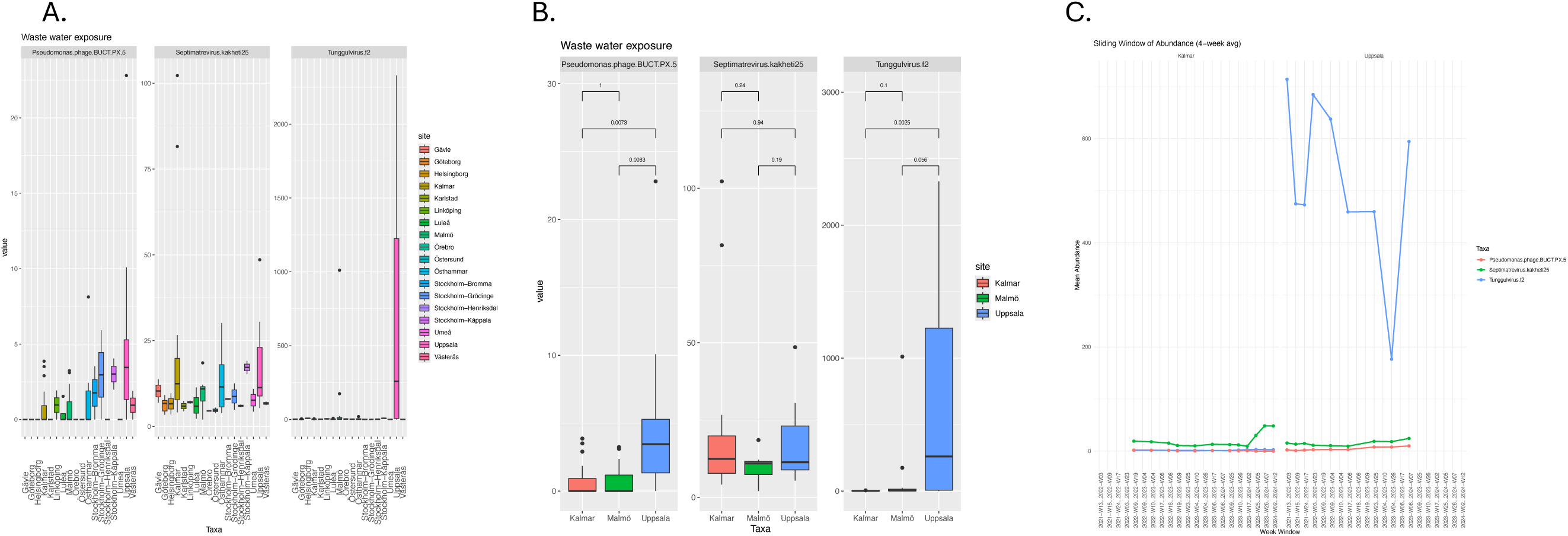
Three prevalent phages infecting *Pseudomonas* bacteria detected in wastewater. **A.** Detection of *Septimatrevirus kakheti25*, *Tunggulvirus f2*, and *Pseudomonas phage BUCT-PX-5* across wastewater samples from 15 municipalities. **B.** Significantly higher cumulative relative abundances of *Tunggulvirus f2* and *Pseudomonas phage BUCT-PX-5* observed in Uppsala compared to Kalmar and Malmö. **C.** Seasonal variation in relative abundance of *Pseudomonas*-infecting phages from summer 2021 to spring 2024 in wastewater from Kalmar and Uppsala.

The human gut *Faecalibacterium species* infecting phages were observed in all wastewater samples (Table 2) across the 15 Swedish municipalities with highly similar relative abundances (Fig 10A). Selecting the Kalmar and Uppsala with most follow-up samples, a significant declining trend in abundance was observed for both phages between 2022 and 2023 and a re-bouncing increase of abundance from 2023 till 2024 (Fig. 10B).

**Figure 10.**
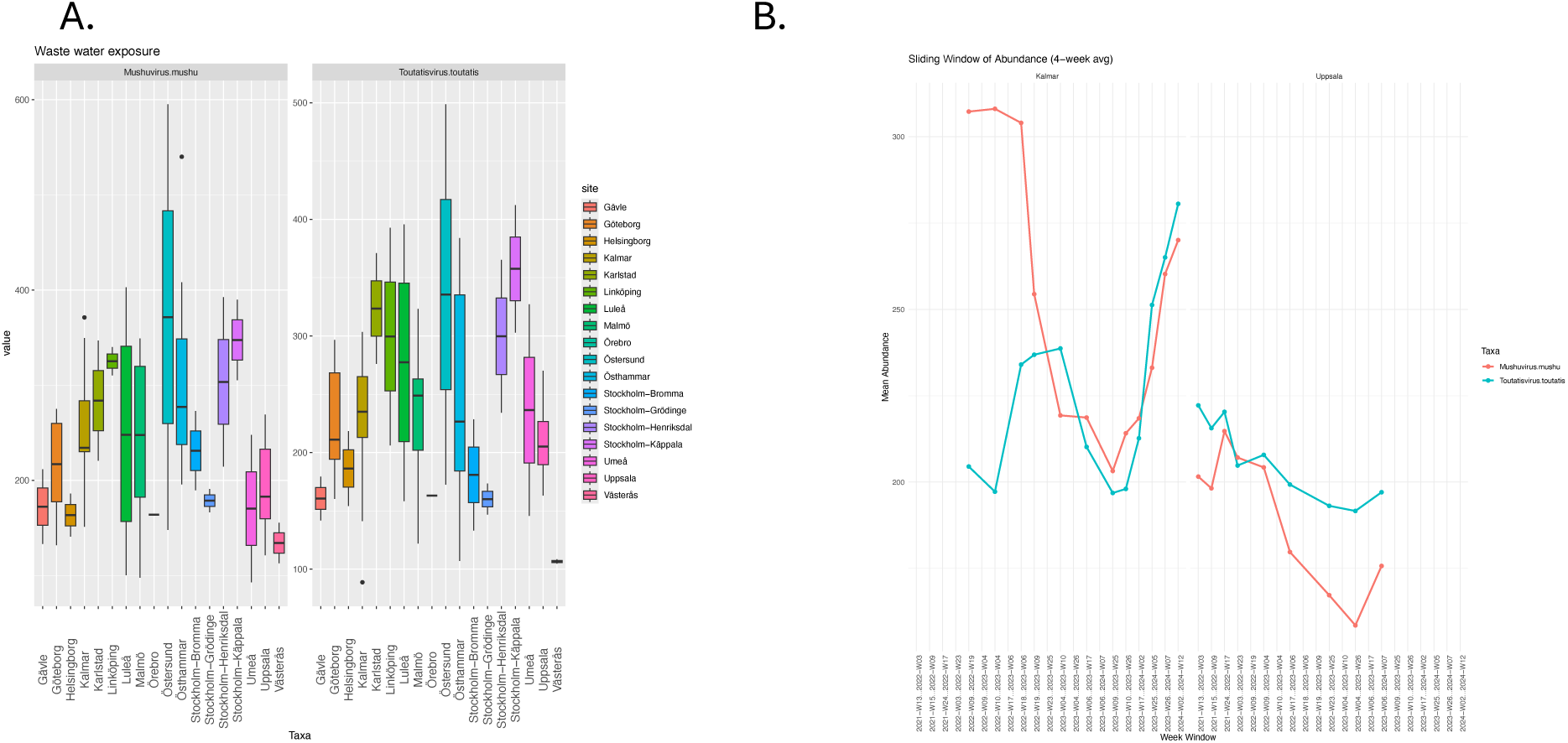
Two prevalent phages infecting *Faecalibacterium* species detected in wastewater. **A.** *Mushuvirus mushu* and *Toutatisvirus toutatis*, phages infecting *Faecalibacterium* spp., were identified across wastewater samples from 15 Swedish municipalities. **B.** Seasonal variation in the relative abundance for both phages was observed between summer 2021 and spring 2024 in wastewater from Kalmar and Uppsala.

The *Salmonella enterica* bacteria infecting phage *Seoulvirus SPN3US*; *Popoffvirus pv56 phage* that infects fish infecting bacteria and *Acinetobacter virus 133 phage* infecting Acinetobacteria were observed in all wastewater samples across the 15 Swedish municipalities (Table 2) but with high fluctuation of the relative abundances between the wastewater origin (Fig 11A). Archaea infecting *Methanoculleus virus L4768 phages* were observed in 46% of the wastewater samples exclusively from Gävle, Umeå and Stockholm-Henriksdal municipalities (Fig 11A). A significant decline in abundance was observed for *Popoffvirus pv56* phage that infects fish infecting bacteria in wastewater from Kalmar between 2022 and 2023 (Fig. 11C).

**Figure 11.**
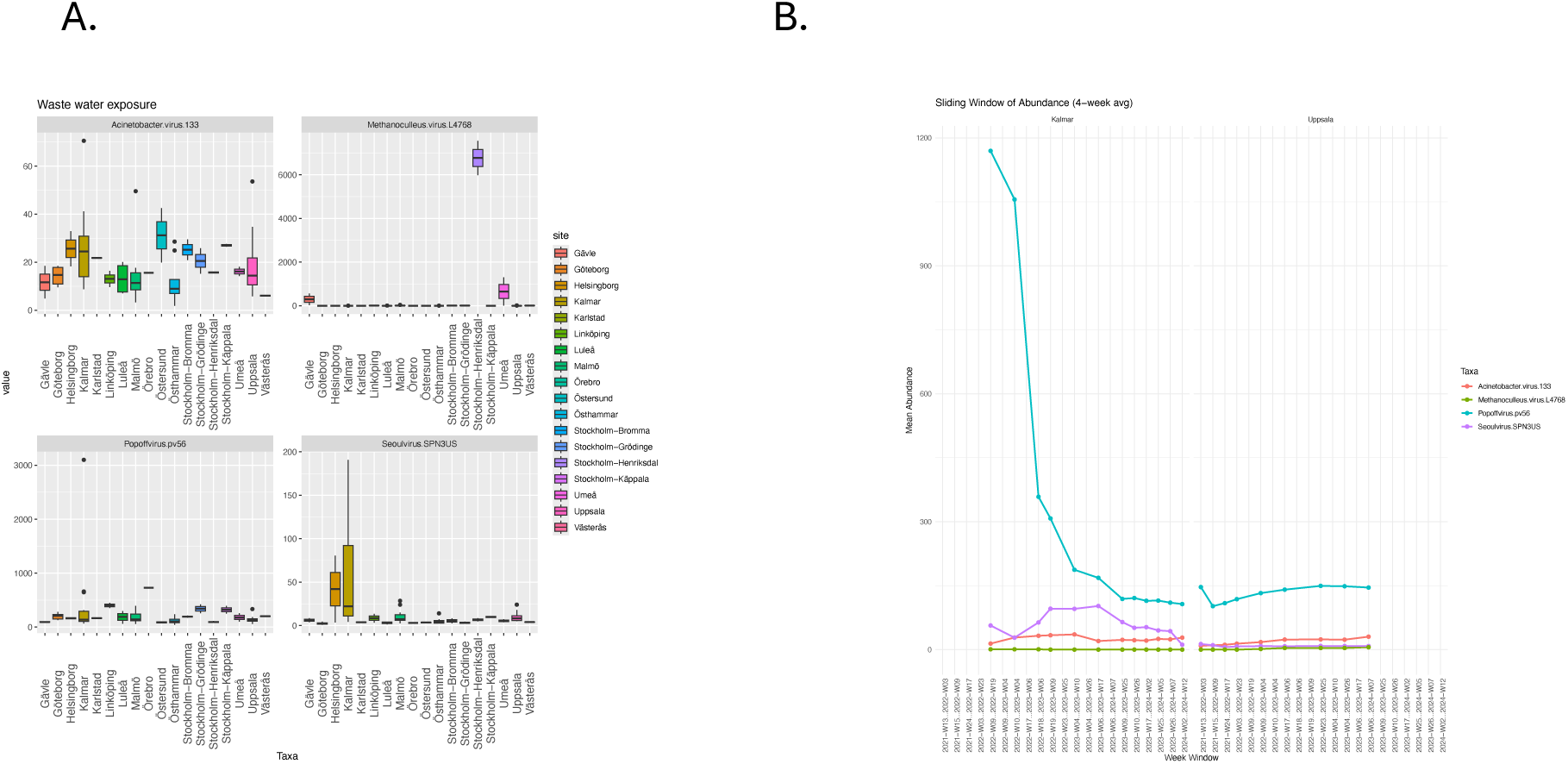
Four additional prevalent bacteriophages detected in wastewater. **A.** *Seoulvirus SPN3US* (infecting *Salmonella enterica*), *Popoffvirus pv56* (infecting fish-pathogenic bacteria), *Acinetobacter virus 133*, and *Methanoculleus virus L4768* were identified across wastewater samples from 15 Swedish municipalities. **B.** Seasonal variation in the relative abundance of *Seoulvirus SPN3US* was observed between summer 2021 and spring 2024 across the three municipalities.

From all the bacterial taxa identified, we selected the fourteen most abundant human-associated bacteria species observed in at least 16% of the wastewater samples for more in-depth regional and temporal analysis (Table 3). Of these, the most abundant bacteria were *Pseudomonas aeruginosa* group, *Acinetobacter baumannii* and *Klebsiella pneumoniae species,* all known opportunistic human pathogens. They were observed in all wastewater samples in this study (Table 3). The composition of the *Pseudomonas aeruginosa* group and *Klebsiella pneumoniae* species varied but not significantly across the different municipalities (Fig. 12A), except for a few outliers (i.e. Västerås and Stockholm – Grödinge). *Acinetobacter baumannii* showed less relative abundance difference across the municipalities (Fig. 12BC). Accounting for the follow-up sampling, Kalmar wastewater revealed a sudden outbreak of *P. aeruginosa* group and *K. pneumoniae* in 2022, followed by a rapid decline of both, while in Uppsala the declining trends for *P. aeruginosa* group and *K. pneumoniae* appeared already between 2022-2023 (Fig. 13A). Similar sudden increases in abundance are shown in 2022 for *Clostridioides difficile, Salmonella enterica* (Fig. 13B), *Shigella flexneri* and *Mycobacterium avium complex* (Fig. 13C).

**Figure 12.**
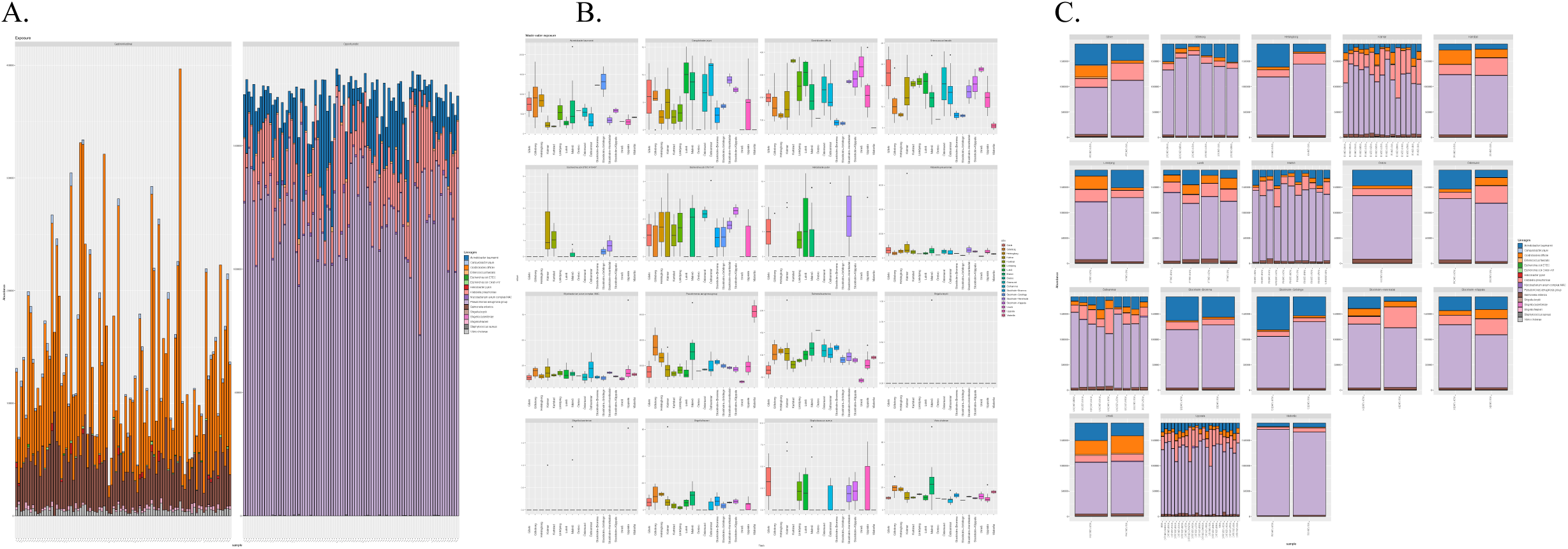
Selected sixteen human pathogenic bacterial species. **A.** Relative proportions of high-quality DNA reads classified into sixteen bacteria pathogens from wastewater samples. **B.** Compositional distribution of the sixteen taxa, and **C.** their regional proportion.

**Figure 13.**
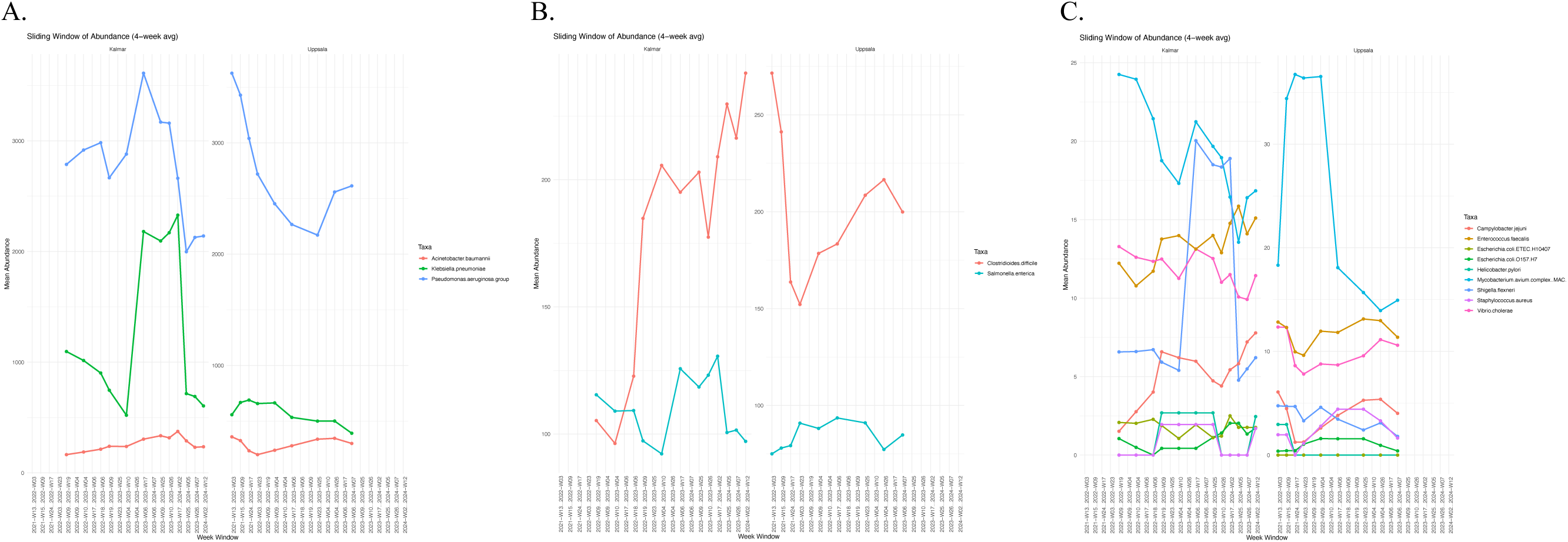
Longitudinal trends of human bacterial pathogens observed in wastewater from Kalmar and Uppsala municipalities between 2021 and 2024. **A.** Sliding window analysis accounting for time of sampling from Kalmar and Uppsala *for P. aeruginosa* group and *K. pneumoniae,* B. for *Clostridioides difficile and Salmonella enterica, and* C. *for Shigella flexneri and Mycobacterium avium complex, Vibrio cholerae, Campylobacter jejuni, Escherichia coli O157:H7, Escherichia coli ETEC H10407, Enterococcus faecalis, Shigella flexneri, Helicobacter pylori and Staphylococcus aureus*.

### Abiotic compounds in wastewater

Our untargeted LC-HRMS analysis of the wastewater revealed an array of 37,122 distinct chemical compounds from which 1,403 were chemicals with known formula and ontology. Several compounds from twenty chemical groups that function as environmental toxins and stressors were observed in the wastewater samples while also a fraction of human activity related chemicals originating from human metabolism as well as from cosmetics, personal care products and from food industry were observed (Table 4). Comparing the chemical wastewater profiles among Kalmar, Malmö and Uppsala - the cities with longitudinal sampling - the 1403 chemicals with known ontology revealed significant relative abundance differences between the three cities (Fig. 14). To further assess longitudinal trends in chemical compositions, we analyzed the relative abundances of chemicals in wastewater positively associated to the AdV F 41 and human gut related *Carjivirus communis* bacteriophages using a four-week sliding window across the sampling period from 2021 to 2024 in Uppsala and Kalmar (Fig. 15). The relative abundance of selected chemicals were significantly higher in Uppsala compared to Kalmar during the period from autumn 2022 to spring 2024 (P_FDR_ < 0.00012), with a local rapid increasing trend observed in Uppsala but not in Kalmar during the monitoring time (Fig. 15). In general, a higher chemical abundances were observed mostly in Uppsala compared to Kalmar during the same time period. A total of 132 significant correlations were observed between biotic and abiotic features mostly related to human activity related compounds (Fig. 16). Among these correlations *Acinetobacter baumannii*, *Carjivirus communis, Carjivirus hominis* and *Kingevirus communis* showed negative correlations while all other microbes showed positive correlations with the chemical compounds (Fig. 17).

**Figure 14.**
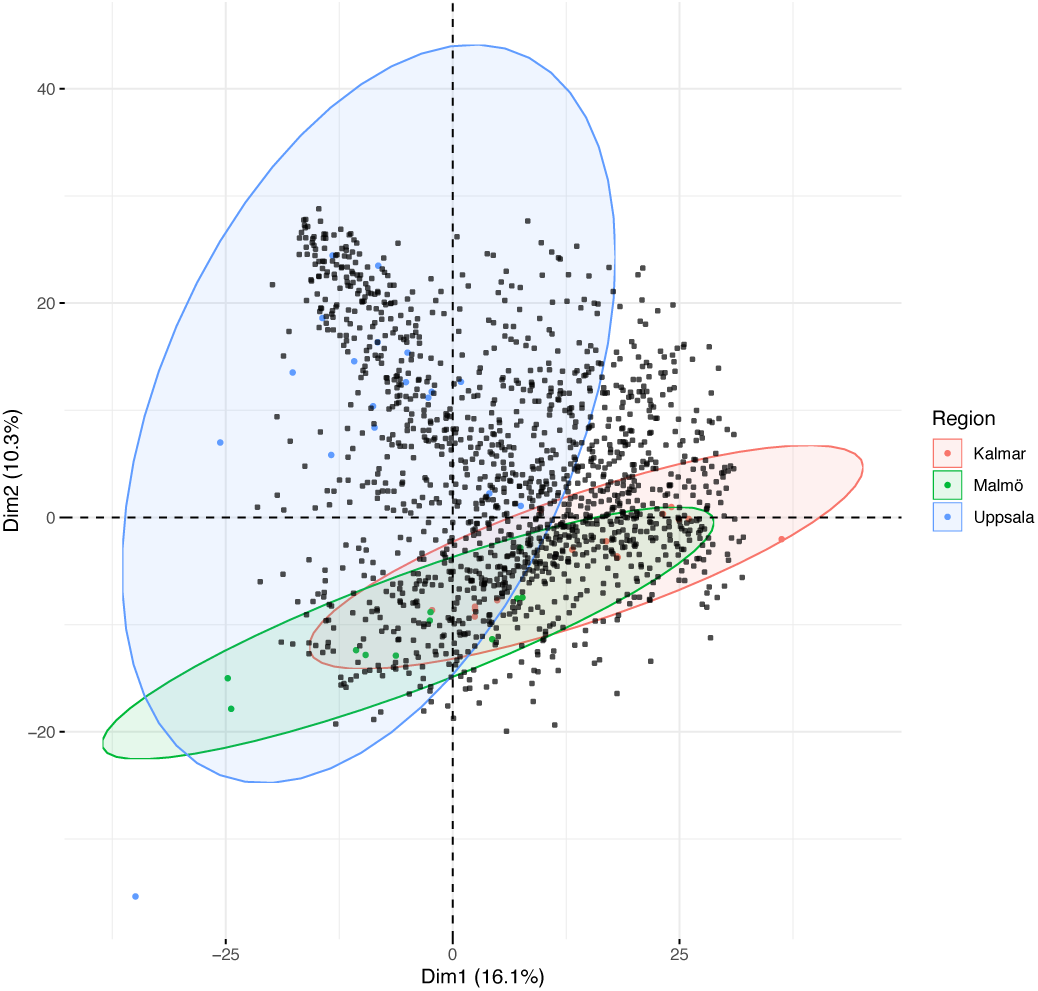
Ordination of all known chemical compounds detected in wastewater. **Significant** (P = 0.001) **c**ompositional difference of 1403 chemicals between the cities of Kalmar, Malmö, and Uppsala.

**Figure 15.**
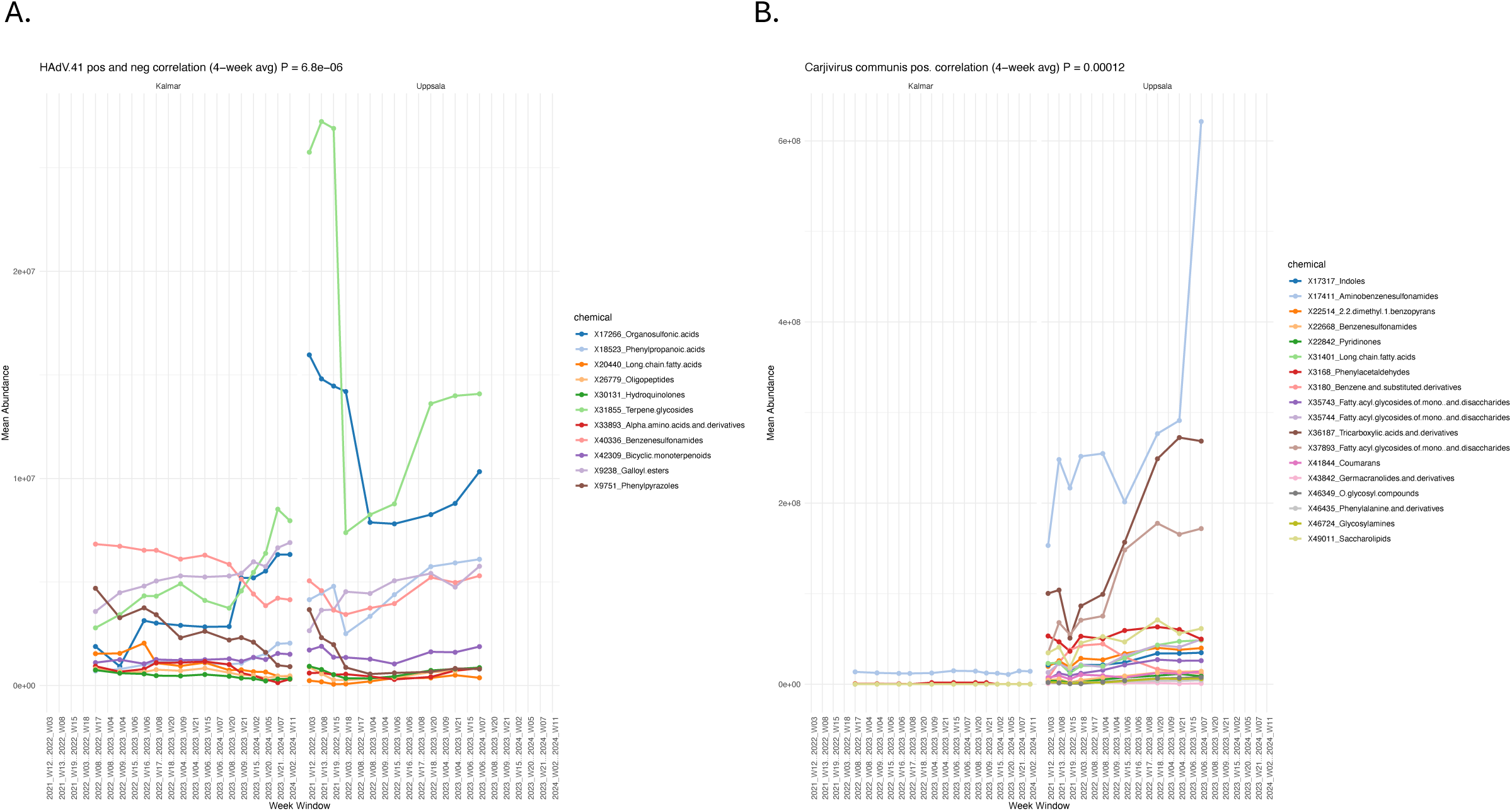
Longitudinal trends of selected chemicals observed in wastewater from Kalmar and Uppsala between 2021 and 2024. A sliding window analysis from Kalmar and Uppsala for chemicals correlated with **A.** Adenovirus F 41 abundances and **B**. correlated with human gut related *Carjivirus communis* bacteriophages abundances.

**Figure 16.**
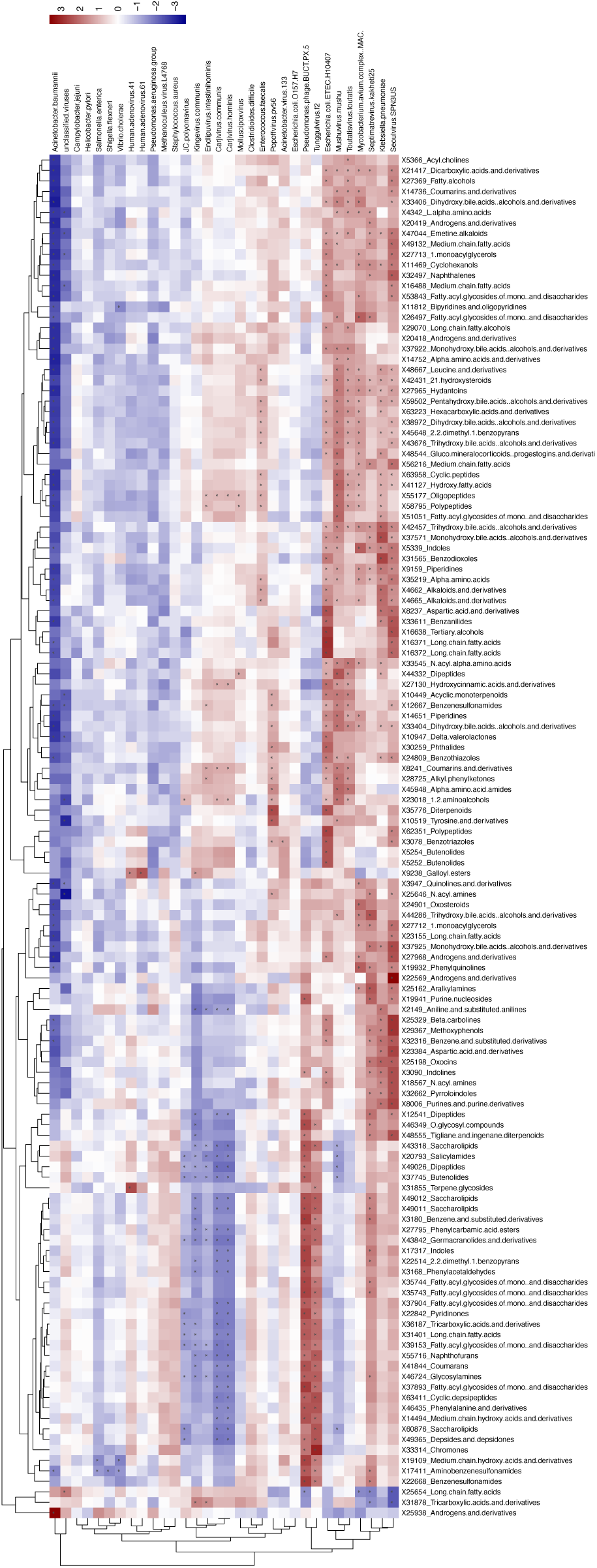
Heatmap of significant biotic and abiotic correlations. Significant positive (red) and negative (blue) correlation of all 1403 chemicals with known ontology with the observed 32 human-related wastewater microbes. Significant correlations are marked with asterix.

**Figure 17.**
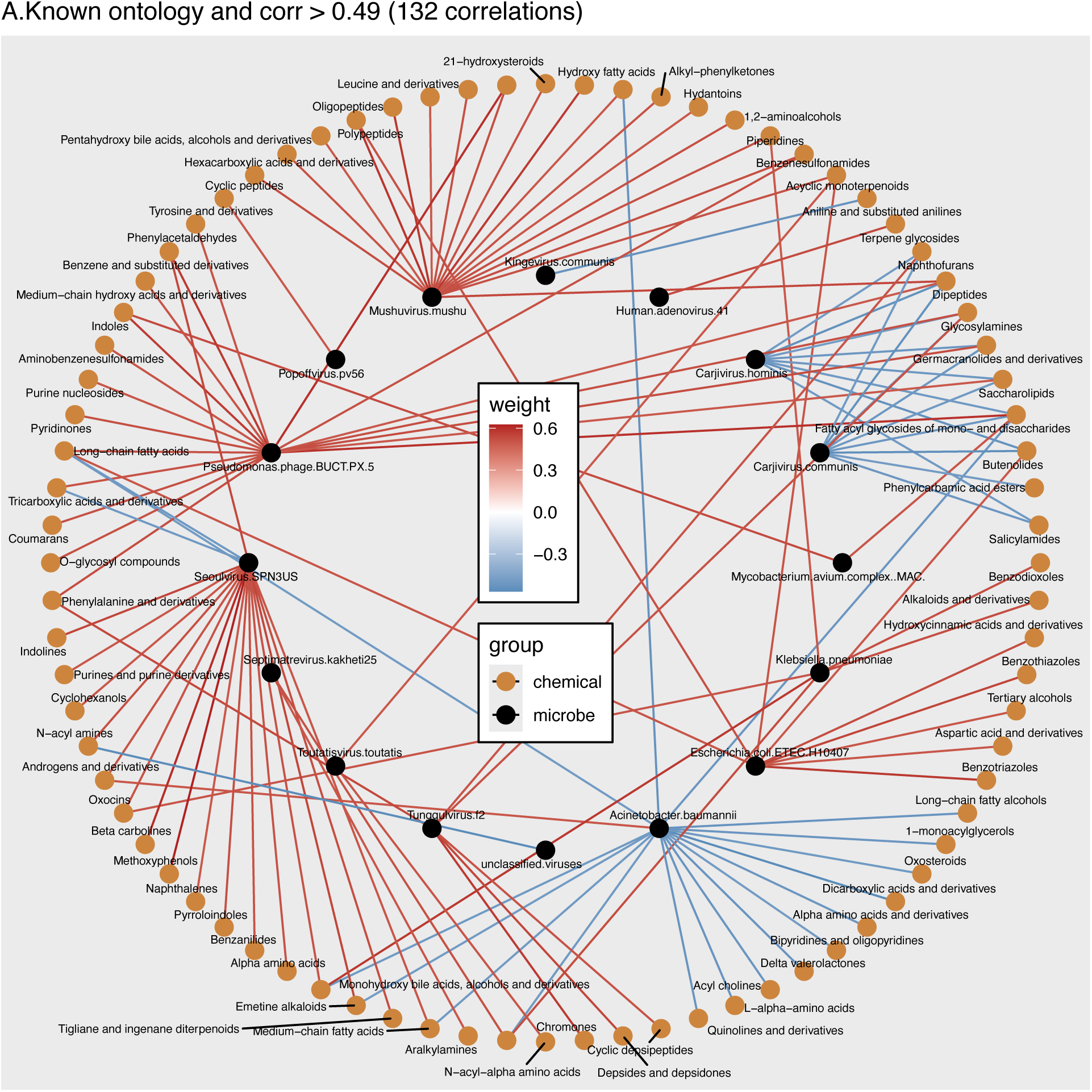
Network of biotic and abiotic correlations. Significant positive (red) and negative (blue) correlation of all chemicals with known ontology and correlation > 0.49 with the observed 32 human-related wastewater microbes.

**Table 4.**
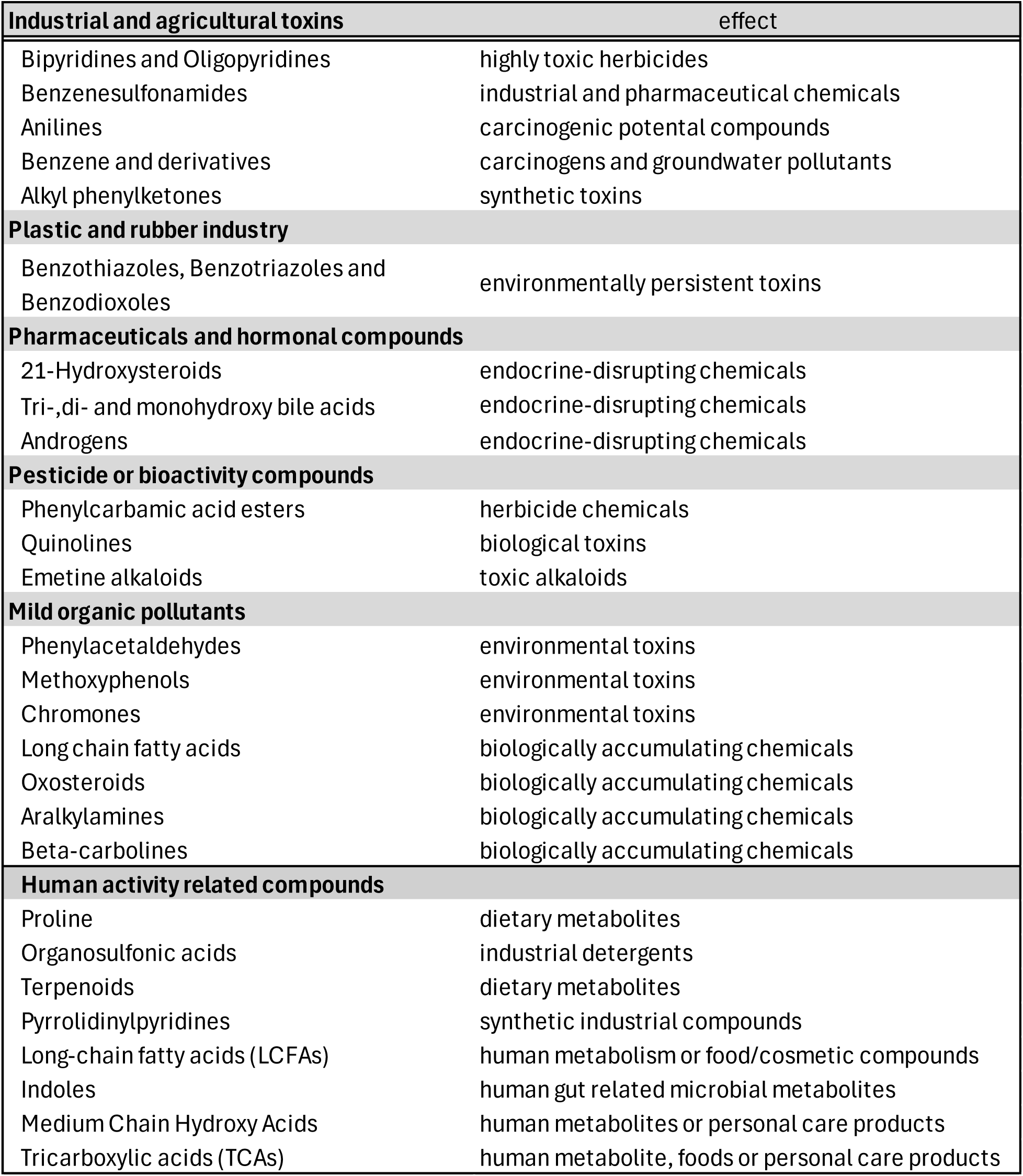
Environmental toxins, stressors and human activity related chemical in wastewater.

## Discussion

The main finding of this study is that biotic and abiotic profiling using shotgun sequencing and untargeted mass spectrometry of wastewater samples enables comprehensive biological and chemical monitoring of not only human bacterial pathogens, but also multiple human viral pathogens and bacteriophages infecting bacteria associated with humans, and microbial interaction with chemicals related to human activities or environmental toxins. From wastewater metagenome sequences, we systematically detected the regional and temporal changes in the prevalence and abundance of viruses and bacteria specific to human health. Most importantly, our findings suggest that social restrictions during the COVID-19 pandemic likely substantially impacted the observed differences in human pathogen prevalence and abundance. In this study, we found that even in rather similar urban environments, the viral relative abundance in wastewater was systematically significantly 1.5-fold higher (P_FDR_=0.000007) in the city of Kalmar compared to Uppsala, both in overall viral abundance and for specific viral lineages during the omicron dominated COVID-19 pandemic in Sweden between summer 2021 till spring 2024. Specifically, we observed a regional outbreak of respiratory infectious diseases causing Adenovirus F41 in Kalmar which associated with the general social relaxation of COVID-19 pandemic restrictions (Fig 7). This is in agreement with a previous wastewater surveillance study in 2022 in Sweden using targeted qPCR and showing local COVID-19 epidemic followed by Adenovirus F41 outbreak in Stockholm (Perez-Zabaleta et al. 2024). Moreover, wastewater monitoring of bacterial pathogens in our study revealed similar trends of local outbreak of *Klebsiella pneumoniae, Clostridioides difficile* and *Shigella flexneri* in Kalmar after the relaxation of pandemic social restrictions (Fig. 13). Other studies have showed seasonal trends of higher viral diversity typically during the winter months and lower diversities in the summer months (Becsei et al. 2024; Smith et al. 2024). We also observed some significant seasonal diversity and distribution patterns in our study. However, the pronounced decline or increase of aerosol-transmitted viral pathogen abundances that we observe is likely caused mostly by the social restrictions and relaxations during and after the COVID-19 pandemia. Similarly, we could trace the likely changes in human activities due to pandemic restrictions and subsequent relaxations from the chemical components in wastewater. These findings highlight the value of wastewater genomic sequencing as a public health surveillance tool with mass spectrometry as a chemical surveillance counterpart, providing insight into the temporal patterns of particular viral populations and chemical exposures within communities and supporting the utility of monitoring infectious disease outbreaks.

Of the six human infecting DNA viruses we detected, the human JC polyomavirus (JCPyV) was the most prevalent and detected in all 91 wastewater samples sequenced in this study. JCPyV is highly prevalent in humans, with 70% to 90% of the adult population showing seroconversion to the virus. Primary infection typically occurs at a young age and leads to a persistent, asymptomatic infection, which may reactivate among immunosuppressed individuals and lead to disease (Padgett et al. 1971). JCPyV is excreted in urine, hence, JCPyV is frequently detected in wastewater worldwide. The viral stability in environmental conditions and resistance to conventional wastewater treatment processes make it a reliable indicator of human fecal contamination. Moreover, phylogenetic studies have identified multiple JCPyV genotypes, which historically present a geographic structure. Hence, wastewater-based surveillance of JCPyV can be used also to trace population movements (Levican et al. 2019).

There are seven known human adenoviruses (HAdVs) species names from A to G with distinct tissue tropism and disease associations. HAdV A, F, and G are linked to gastrointestinal disease, while HAdV B, C and E cause respiratory infections, some particularly common in healthcare environments. While most HAdV infections are generally mild, these viruses are highly prevalent and can cause serious illnesses in vulnerable populations. Particularly, HAdV F 41 serotype is a leading cause of pediatric gastroenteritis, and it has been recently also associated with acute hepatitis of unknown etiology by WHO (WHO 2022). In our study we systematically find HAdV F41 in 93% of the Swedish wastewater samples and throughout the follow-up between 2020 and 2024. Moreover, we observed a low abundance of the HAdV F41 during the pandemic and a subsequent regional outbreak of HAdV F41 in Kalmar, likely associated with the social relaxation of COVID-19 pandemic restrictions (Fig 7).

### Gut bacteriophages

The detection of bacteriophages such as *Carjivirus communis, Carjivirus hominis, Endlipuvirus intestinihominis* and *Kingevirus communis* in wastewater provides important insights into the structure and dynamics of the human gut virome and highlights the growing utility of WBS for monitoring population-level health and microbial ecology. These phages belong to the order Crassvirales, a diverse group of double-stranded DNA viruses that are among the most abundant and widespread components of the human gut microbiome, particularly infecting members of the Bacteroides. Their prevalence in fecal metagenomes across populations worldwide underscores their global ecological significance and highlights their potential as biomarkers for human fecal contamination in environmental waters. *Carjivirus communis,* often referred to as the prototypical crAssphage contains elements typical of both phages and plasmids, including toxin-antitoxin modules, and mobilization genes. These features suggest a hybrid lifestyle, allowing for long-term persistence in host bacteria and complex host-phage interactions. *Carjivirus hominis* shares similar genomic architecture, supporting the concept of extrachromosomal maintenance and possibly co-evolution with human-associated Bacteroides. Other members, such as *Endlipuvirus intestinihominis* and *Kingevirus communis* exhibit substantial phylogenomic divergence with no clear core genome that reflects the adaptive specialization of these phages to distinct host lineages within human gut ecosystems.

*Mushuvirus mushu* and *Toutatisvirus toutatis* recently identified through metagenomic surveillance expand the recognized diversity within Crassvirales and reflect the capacity of wastewater surveillance to uncover novel, uncultivated phage taxa. While functional roles of many crAss-like phages remain unresolved, their recurring detection in untreated and treated wastewater not only mirrors human gut microbial excretion but also positions them as non-invasive indicators of community health, gut microbial shifts, and sanitation effectiveness. However, recent global metagenomic analyses indicate that only a subset of Crassvirales demonstrate high human specificity, and this specificity does not correlate with shared evolutionary origin but rather reflects independent adaptation events to human-associated Bacteroides strains. Moreover, these human-specific phages exhibit heterogeneous geographic prevalence, shaped by regional differences in host bacterial populations, diet, and hygiene infrastructure. The presence of human gut related phages in wastewater offers a scalable approach to tracking human fecal inputs, microbial dynamics, and potentially gut health at the population level. These phages stand out as promising targets for public health monitoring, environmental virology, and advancing our understanding of host-microbe-virus interactions in the human microbiome.

### Human related wastewater bacteria

The detection of a diverse array of clinically relevant bacterial taxa in wastewater highlights the utility of WBS for public health monitoring, particularly for tracking antimicrobial resistance and enteric disease risks. We identified pathogens such as *Klebsiella pneumoniae, Escherichia coli ETEC H10407* and *Salmonella enterica* each known for causing severe gastrointestinal and systemic infections. The presence of *K. pneumoniae*, a gut-resident and major cause of healthcare-associated infections, is especially concerning given its capacity for multidrug resistance, posing risks of environmental dissemination from hospital sources. Similarly, the detection of *E. coli ETEC*, a leading cause of traveler’s and childhood diarrhea, and *S. enterica*, a major foodborne pathogen, indicates ongoing fecal contamination and potential transmission through food and wastewater.

These findings affirm that wastewater sustains a complex mixture of pathogens reflective of community-wide infection burdens. Their continuous monitoring provides actionable insights into population health, disease emergence, and the circulation of antimicrobial-resistant organisms, thus reinforcing the value of WBS as a non-invasive, real-time epidemiological approach for public health monitoring. Wastewater is a reservoir and surveillance matrix for both bacterial pathogens and their viral counterparts. Phages mirror the microbial composition of the human microbiome but also offers potential as biomarkers of pathogen presence and as biocontrol agents for mitigating antibiotic resistance in wastewater treatment systems. Moreover, their detection with environmental chemicals can help assess the selective pressures imposed by antibiotics, disinfectants, and other environmental stressors prevalent in wastewater, which shape both phage-host dynamics and resistance gene flow. Together, these findings reinforce the utility of WBS for monitoring pathogen-phage networks in real-world settings, providing critical data to inform outbreak detection and public health interventions.

Taken together, systematic and integrated viral, bacterial, and chemical wastewater exposomics provides a comprehensive framework for monitoring population-level environmental exposures. By capturing both known and unclassified biological and chemical signatures, this approach enables high-resolution mapping of dynamic changes in the human environmental exposome at community-level. It offers a non-invasive and scalable means to detect shifts associated with behavioral, seasonal, or policy-driven factors, such as pandemic-related interventions. The integration of multi-omic data supports the early identification of local outbreak signals and emerging threats. Ultimately, wastewater exposomics serves as a critical tool for real-time public health surveillance and environmental risk assessment.

## Data Availability

All data produced in the present study are available upon reasonable request to the corresponding author.

https://github.com/PimenoffV/

## Acknowledgements

This work correspond to specific aims of grant 874662 to the HEAP consortium from the European Union for creating the wastewater biotic results supported by the Finnish Functional Genomics Center services in Finland and abiotic results supported by the SciLifeLab National Facility for Exposomics (Metabolomics and Exposomics Platform), Stockholm, Sweden.

